## Supporting information for "How Informative Were Early SARS-CoV-2 Treatment and Prevention Trials? A longitudinal cohort analysis of trials registered on clinicaltrials.gov" (from another version of this preprint: version 1, 2021-09-05)

S1 File. Trial Inclusion and Exclusion Criteria

S2 File. Data downloaded from ClinicalTrials.gov

S3 File. Additional data points

S4 File. Assessment of Trial Redundancy

S5 File. Protocol Deviations

S1 Figure. PRISMA Diagram

S2 Figure. Ratio of Actual to Estimated Number of Patients Enrolled

S3 Figure. Ratio of Actual to Estimated Recruitment Length

**S1 File. Trial Inclusion and Exclusion Criteria**

Inclusion criteria:

- Interventional clinical trials
- Trial Status: Completed, Terminated, Suspended, Active not recruiting, Enrolling by invitation, Recruiting
- Trials testing an efficacy hypothesis in the primary outcome
- Phase numbers: Phase 1/2, Phase 2, Phase 2/3, Phase 3
- Drug, biological, surgical, radiotherapy, procedure and device interventions

Exclusion criteria:

- Behavioral interventions (including psychological, dietary, physiotherapy, etc.)
- Trials of natural products (not regulated by the FDA)
- Phase 1 trials (do not typically evaluate efficacy outcomes and are not required by FDA to register)

**S2 File. Data downloaded from ClinicalTrials.gov**

- NCT Number
- Link to ClinicalTrials.gov record
- Title of Trial
- Indication
- Intervention
- Phase
- Enrollment (current actual or anticipated enrollment and original estimated enrollment)
- Age
- Gender
- Trial status
- Reason for termination/suspension (for trials that are terminated or suspended)
- Allocation (randomization)
- Interventional model (Single group versus multiple group assignment)
- Masking
- Trial Start date (current start date and original start date)
- Trial projected close date
- Lead sponsor
- Funder type
- Number of arms
- Study location (country location of each center participating in the trial)
- Primary outcome

**S3 File. Additional data points**

Data points requiring human curation were independently extracted by two individuals using Numbat software – an open source tool created by BGC, available at https://numbat.bgcarlisle.com.

This included the following items:

- Categorization of trial type (SARS-CoV-2 Treatment trial versus Prevention trial versus unclear/not stated).
    - A SARS-CoV-2 treatment trial was defined as a clinical trial conducted in a population with documented SARS-CoV-2 infection designed to evaluate the safety and/or efficacy of an intervention directed at improving clinical or surrogate outcomes related to the disease.
    - A SARS-CoV-2 prevention trial was defined as a clinical trial conducted in a population without known SARS-CoV-2 infection designed to evaluate the safety and/or efficacy of an intervention directed at preventing serologic conversion and/or clinical disease due to SARS-CoV-2. These studies are often conducted in an at-risk population, whereby exposure to SARS-CoV-2 is felt to be likely. They can include populations who have been exposed to SARS-CoV-2 (post-exposure prophylaxis), without documented infection or clinical symptoms of disease, in addition to populations who may be exposed to SARS-CoV-2 at a future time (pre-exposure prophylaxis).

- Location of care (Ambulatory versus Hospitalized versus Intensive Care versus unclear/not stated)

- Illness severity (Healthy patient versus Asymptomatic versus Mild versus Moderate versus Severe versus Critical versus unclear/not stated)
    - Illness severity was as stated by investigators, or, if this was not provided, it was inferred using the WHO COVID-19 disease severity classification.[1]

- Treatment type (guided by the World Health Organization (WHO) COVID Classification of treatment types[2]

- Presence of a comparator trial arm

- Presence of a placebo or standard of care trial arm

- Primary outcome classification (Clinical outcome versus Surrogate outcome versus Procedural outcome versus Test result (the latter was designated for prevention trials)
    - A clinical outcome is one that specifically evaluate quality of life, morbidity/disability or mortality. Examples of COVID-19 clinical outcomes

from[3]: i) all-cause mortality; ii) respiratory failure (including need for mechanical ventilation, ECMO, non-invasive ventilation, high-flow nasal cannula); iii) need for intensive care unit level care; iv) need for hospitalization; v) objective measures of sustained improvement (e.g. return to baseline oxygen requirement); vi) sustained clinical recovery. Clinical outcomes may be presented on their own (as a single clinical outcome), as a composite outcome, or as an ordinal scale combining multiple clinical outcomes.

- A surrogate outcome is one that does not directly measure the impact of an intervention on quality of life, morbidity/disability or mortality. Rather, it is designed to predict a clinical outcome. For example, virologic response (virologic clearance / viral load) is an example of a COVID-19 surrogate.

- Inclusion of elderly patients (aged 60 years and above)

- For terminated trials, classification of reason for termination (based on that stated in the "why stopped?" field in the trial registration record (Feasibility versus Efficacy/Safety/Progress of Science versus unclear/not stated)

**S4 File. Assessment of Trial Redundancy**

Assessment of trial redundancy included both automated and human-curated steps. Using datapoints directly downloaded from ClinicalTrials.gov and those previously independently assessed by KK & LZ, an automated comparison of trial phase, type of trial (treatment versus prevention), location of care, illness severity, age (binary assessment of inclusion/exclusion of population aged $\geq$ 60), and treatment regimen (using human-curated synonyms for interventions) was carried out using R version 3.6.3. Trials with a direct match on all the above elements were advanced to independent assessment of trial primary outcome, comparator arms and other (including type of population (for example, healthcare workers) and trial aim (for example, pre-exposure prophylaxis versus post-exposure prophylaxis) by two authors (NH & KK).

Trials were classified as redundant if there was a comparator trial with an earlier start date of the same phase, location of care, severity, age, treatment regimen, primary outcome (primary outcome of the earlier comparator trial had to at least encompass that of the trial of interest), treatment arms (arms of the earlier comparator trial had to at least match those in the trial of interest), population and aim.

**S5 File. Protocol Deviations**

1. We redefined the terms for our measures of effect as follows:

- “Importance” as “Potential redundancy”,
- “Feasibility” as “Feasibility of Patient-Participant Recruitment”.

2. We calculated the proportion of trials meeting all 3 criteria of informativeness.

3. We performed a *post hoc* assessment of trial similarity, based on assessment of trial type, phase, patient-participant characteristics and treatment regimen.

4. We will be publishing the results of our analysis of feasibility of recruitment by location in a separate manuscript.

**S1 Figure. PRISMA Diagram**

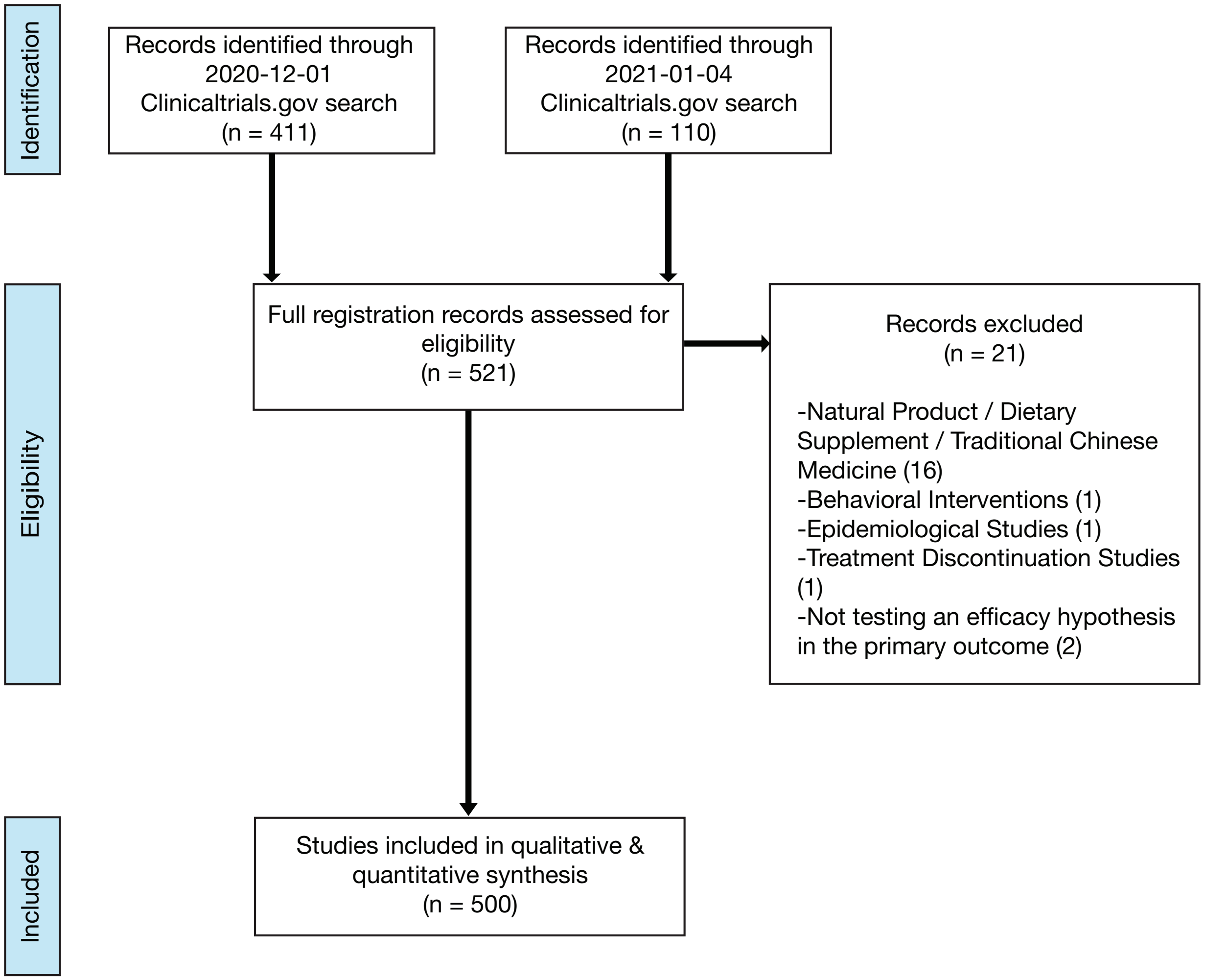

**S2 Figure. Ratio of Actual to Estimated Number of Patients Enrolled**

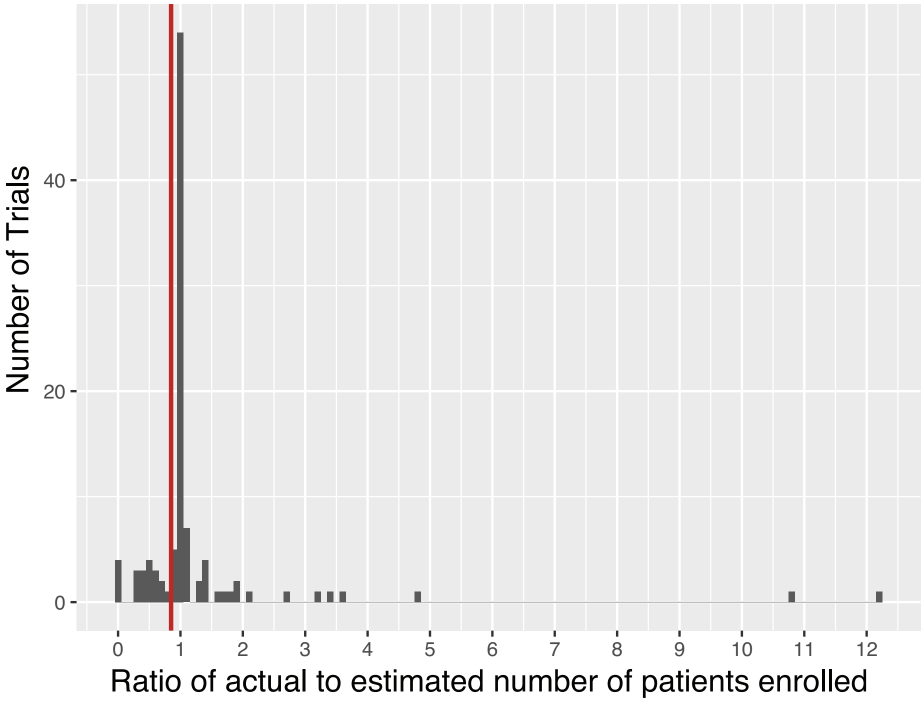


Vertical red line indicates 85% goal enrollment (threshold for recruitment feasibility)

**S3 Figure. Ratio of Actual to Estimated Recruitment Length**

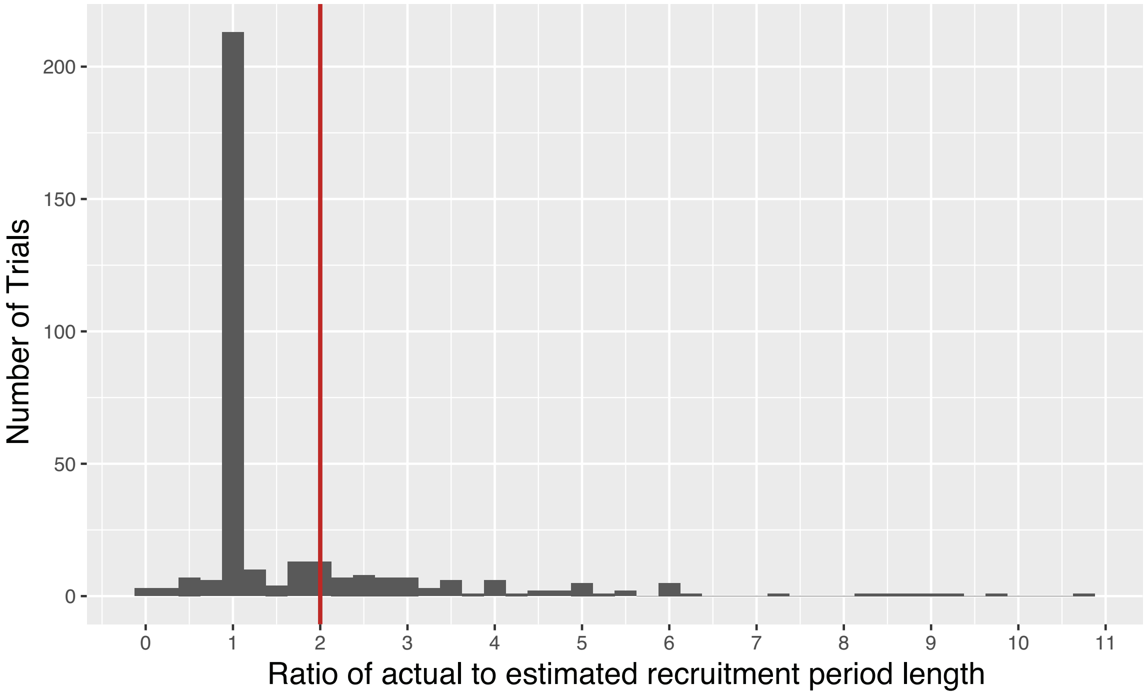


Vertical red line indicates two times the intended recruitment length
